# SARS-CoV-2 variants of concern are associated with lower RT-PCR amplification cycles between January and March 2021 in France

**DOI:** 10.1101/2021.03.19.21253971

**Authors:** Benedicte Roquebert, Stéphanie Haim-Boukobza, Sabine Trombert-Paolantoni, Emmanuel Lecorche, Laura Verdurme, Vincent Foulongne, Sonia Burrel, Samuel Alizon, Mircea T. Sofonea

## Abstract

SARS-CoV-2 variants raise concern regarding the mortality caused by COVID-19 epidemics. We analyse 88,375 cycle amplification (Ct) values from variant-specific RT-PCR tests performed between January 26 and March 13, 2021. We estimate that on March 12, nearly 85% of the infections were caused by the V1 variant and that its transmission advantage over wild type strains was between 38 and 44%. We also find that tests positive for V1 and V2/V3 variants exhibit significantly lower cycle threshold (Ct) values.

## 1 Context

At least three SARS-CoV-2 lineages are currently a major source of concern: variant V1 from lineage B.1.1.7 [1, 2], variant V2 from lineage B.1.351 [3], and variant V3 from lineage P.1 [4]. V1 and V3 variants have been shown to be more contagious [1, 2, 4], while V2 and V3 variants seem to evade immune responses [3, 4]. Although the mechanistic bases are still being investigated, the increased contagiousness could be driven by the N501Y mutation and the Δ69-70 deletion in the Spike protein [1]. The immune escape could largely be due to the E484K mutation also in Spike [3, 4]. Current evidence regarding potential differences in cycle threshold values (denoted Ct) are still unclear [5, 4] as how well they reflect viral loads [6, 7].

In France, using data from 11,916 tests performed on Jan 6 and 7, 2021, a study estimated that 3.3% of the infections were caused by the V1 variant at that time [8]. With data from 10,261 performed on Jan 27, 2021, their estimate increased to 13.0%. Another study used 40,777 tests performed between Jan 26 and Feb 16, 2021, and estimated that on Feb 16, 55% of the infections were caused by V1, V2, or V3 variants [9].

Here, we estimate the spread of SARS-CoV-2 variants of concern using variant-specific RT-PCRs performed in nasopharyngeal (NP) swabs from the general population and in hospital. Furthermore, we analyse the Ct values of these tests.

## 2 Dataset

We analyse 105,356 variant-specific RT-PCR tests performed in France on the same number of individuals between Jan 26 and Mar 12, 2021. The main assay used was ID™ SARS-CoV-2/UK/SA Variant Triplex (ID SOLUTION) but for 3,901 tests (3,7%) performed before Feb 2, 2021 we used the VirSNiP SARS-CoV-2 Spike del+501 (TIB MOLBIOL) assay. The sampling varied between French regions and we excluded from the analysis regions with less than 200 tests. 934 tests were also removed because the sampling region was missing.

These tests have probes with 3 targets: a control one in the virus N gene, the Δ69-70 deletion, and the N501Y mutation. For V1 variants, both the deletion and the mutation are present. For V2 or V3 variants, only the N501Y is detected. For VirSNiP assay, it is based on 501 and 69/70 fragments amplified and analyzed with a melting curve using mutation-specific probes, as described earlier [9]. As indicated in [9], the test specificity was confirmed internally using next-generation sequencing.

The main cofactors in the analysis were the assay used, the patient age, the sampling date, the sampling region, and the sampling facility (hospitals or city screening).

For 88,375 ID SOLUTION tests, we also analyse the cycle threshold value (Ct) of the virus control gene of the assay. Ct values greater than 30 were ignored because they may provide unreliable results regarding the variant-specific probes LoD (Limit of Detection). Indeed, the latter are located in the S gene, which tends to exhibit higher Ct values than the N gene [10].

## 3 V1 variant is dominant

Using the same methodology described in [9] and in Appendix, we calculated the transmission advantage of each variant compared to the wild type strain after correction for several biases (region, sampling date, assay, and patient age). This inference was performed without the data from hospitals (in order to avoid a sampling delay bias) and for individuals from 5 to 80 years old. For V1, the increase in transmission was 41% (95% confidence interval, CI: [38,44]%). For V2-V3, the estimate was 27% (95%CI: [25,29]%). These results assume the serial interval reported in [11].

Based on these inferences, we estimated the proportion of new infections caused by each type of strain on Mar 12, 2021. At a national level, the estimate was 84,6% for V1 and 5,4% for V2/V3, but with strong regional heterogeneity (Figure 1).

**Figure 1:**
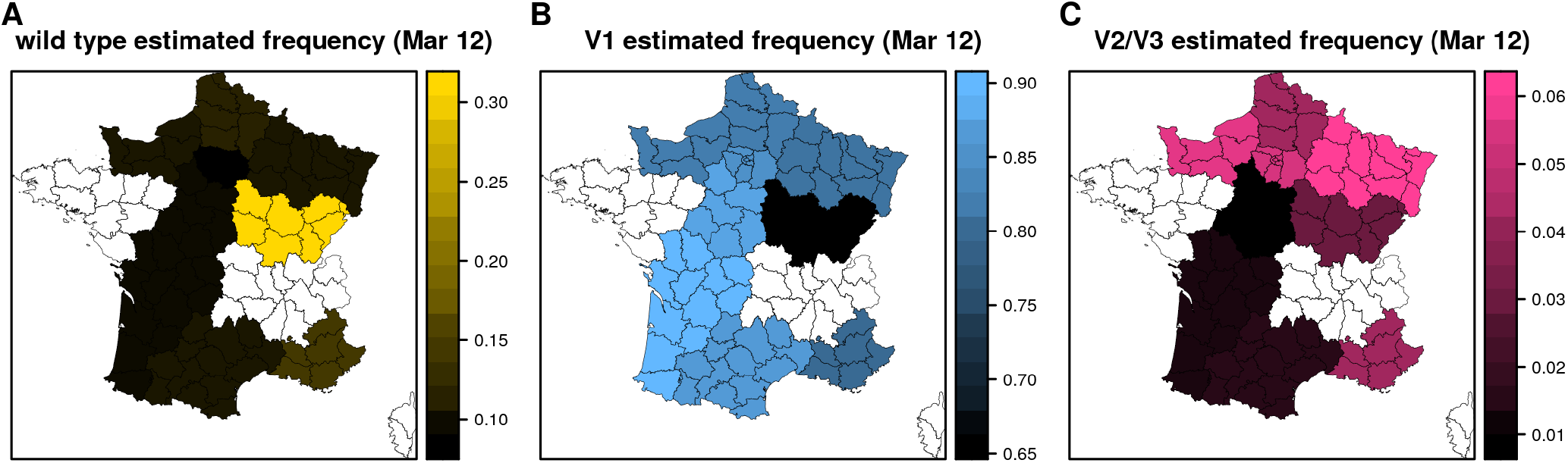
Estimated proportion of new infections caused by A) wild type, B) V1 variant, and C) V2 or V3 variants on Mar 12, 2021, in French regions. Regions with insufficient sampling are in white.

## 4 Variants have lower Ct values

We analysed tests from patients from 1 to 89 years old with Ct values lower than 30. We excluded tests where the strain could not be determined. Overall, this represents 63,924 tests (96% of all the tests with Ct values).

We used a multivariate linear regression to study variations in Ct values between strains. The linear model covariates were the age, the sampling facility (hospital or city), the sampling date, and the region. We also considered an interaction between sampling region and date. We used a type-I analysis of variance (ANOVA) and added the variant covariate last to avoid favouring it in the analysis.

The model residuals were normally distributed and a likelihood-ratio test between a model with or without strain effect retained the former. Overall, the model explained a small fraction of the variation in Ct (adj. R^2^=4.4%), which is consistent with these values being highly variable [10].

The virus strain effect was highly significant in the ANOVA (Figure 2). Samples from V1 variants had a significantly smaller Ct than that from V2/V3 variants (21.9 vs. 22.2). Both had significantly smaller Ct values than wild type strains (23) and other variants (without the N501Y mutation but with the Δ69-70 deletion).

**Figure 2:**
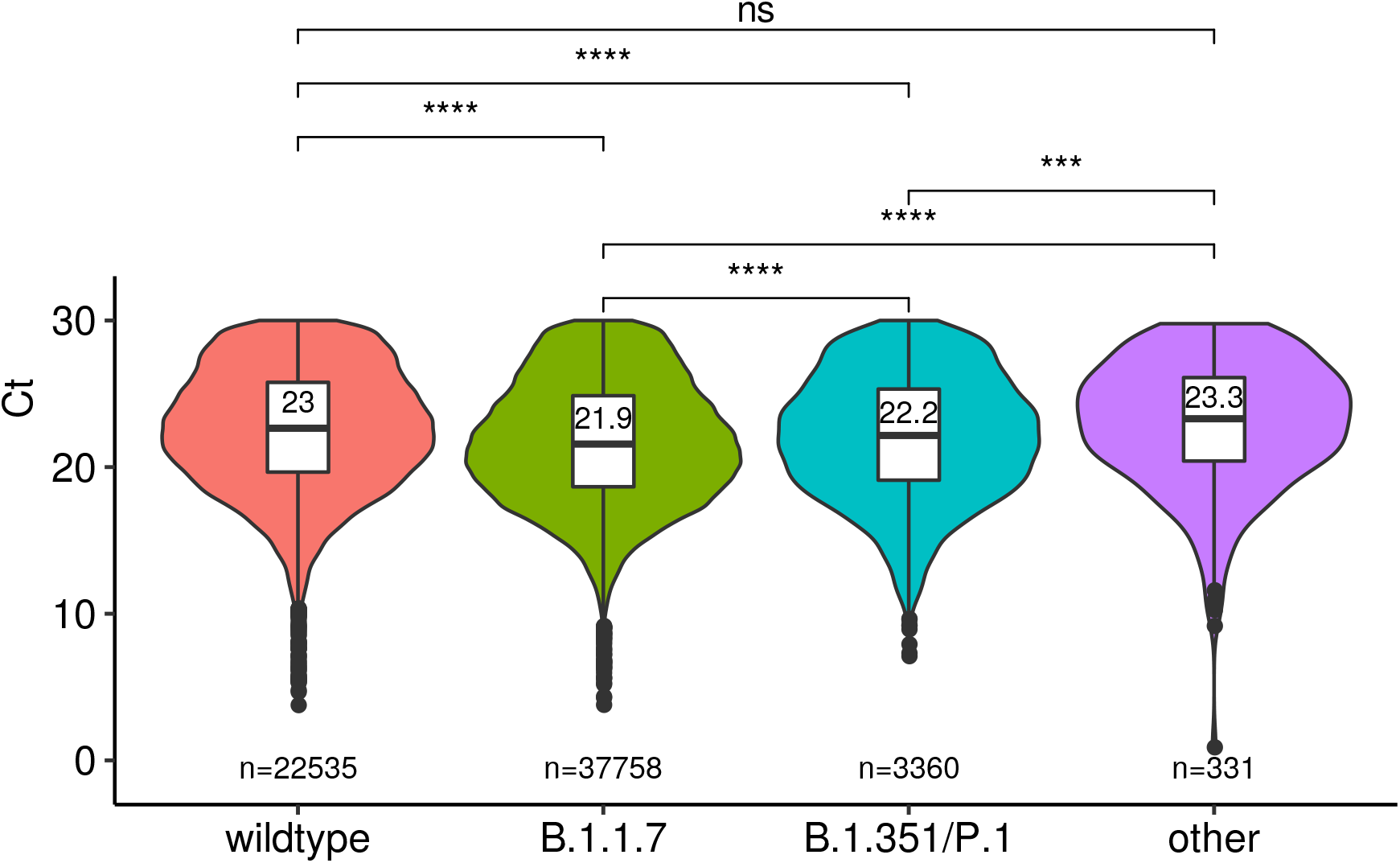
Cycles threshold (Ct) value for SARS-CoV-2 strains. Median estimates based on the linear model are shown in the box plot, and number of tests in each class are shown in the bottom of the graph. ‘other’ indicate tests with the Δ69-70 deletion and without the N501Y mutation. Start indicate the significance level (∗ ∗ ∗ ∗ for a p-values strictly lower than 10^*−*4^ and ∗ ∗ ∗ for a p-value of 10^*−*4^, ns: not significatif).

The model also indicated a significant decrease of Ct with age, which is also consistent with existing data [10]. The sampling date was non-significant but the sampling region mattered and so did the interaction between sampling date and region. Samples from hospitals has a slightly higher Ct, but this is likely due to the fact that testing in the general population occurs approximately 7 days after infection, whereas hospital admissions occur after a median time of 14 days [12]. Therefore, data from hospitalised patients is likely to reflect an older state of the epidemic.

## 5 Discussion

We show that variant of concern V1 is now vastly dominant in France compared to wild type strains (84,6% vs. less than 10%). V2 or V3 variants remain limited (approximately 5.4% of the new infections). These results are consistent with earlier reports of a marked transmission-advantage of the V1 variant [1, 2, 9, 8].

By investigating the RT-PCR Ct values, which can inform us on clinical features of the infection [6, 10], we show that infections caused by variants significantly differ from that caused by wild type strains. That variants are associated with lower Ct values could be an indication of higher viral load, although care must be taken because of the biology of SARS-CoV-2 [7] and of the variability inherent to such values [10].

This result contrasts with earlier findings. One study did not find a significant result when comparing Ct values for tests with or without the S-gene target failure [5]. However, our results are based on a variant-specific PCR. Another study on the V3 variant [4] did not find a significant difference after accounting for the symptom onset to sampling delay. However, their study was performed on a limited number of samples (*n* = 147).

## Data Availability

The data and scripts used for the analysis will be shared upon peer-reviewed publication.

## Data availability

The data and scripts used for the analysis will be shared upon publication.

## Acknowledgements

We thank the ETE modelling team for discussion, as well as the CNRS, the IRD, the ANR, and the Région Occitanie for funding (PHYEPI grant).

This study was approved by the Internal Review Board of the CHU of Montpellier (ClinicalTrial.gov identier NCT04738331).

## Supplementary methods

### Linear model for the Ct analysis

We used a type I error for the analysis-of-variance. Our response variable was the Ct value. The main covariate of interest was the strain and it could take 4 values (V1, V2 or V3, wild type, or other). The other covariates were the age, the sampling facility (hospital or city), the sampling date, and the geographical region. We also considered an interaction between sampling region and date. We used a type-I analysis of variance (ANOVA) and added the strain covariate last. The motivation for this is that with the sequential assumption of the summing of the squares (type I method), the order in which the covariates are tested matters, and, in the case of an uneven sampling, the last one in the list is less likely to be significant. Therefore, our assumption decreases the risk of erroneously attributing observed variance to a variant effect.

We used a likelihood-ratio test to determine whether the addition of the strain effect statistically improved the explanation of the data.

### Generalised linear model to correct for variant sampling bias

As indicated in [9], for a given variant category (V1 or V2/V3) we first perform a generalised linear model with a binomial error distribution where the variable of interest is the binary variant variable (with values ‘variant’ or ‘wild type’) and the explanatory variables are the sampling date, the sampling region, and the individual age. We also include an interaction between sampling region and date. We then use the residuals of this model to infer the transmission advantage of the variant.

### Logistic growth fitting

We used the fitted values of a GLM model applied to the data after removing samples from hospitals (the sampling location effect was also obviously removed from the model) to perform the inference of a two-parameter logistic growth kinetic curve: *f* (*t*) = (1 + e^*−ρ*(*t−τ*)^)^*−*1^, where *f* (*t*) is the frequency of the variants in the new infections at time *t, ρ* is the relative growth rate of the variants and *τ* is the time at which *f* reaches 1*/*2. This method is indeed more appropriate to deal with temporal auto-correlation biases in proportion time series [1, 2].

The parameter estimation was performed using the drc package in R both at the national and the regional level (for regions with at least 1,000 samples). The confidence intervals of the fitted curves rely on those of the estimated date of reaching half proportion of new infections (*τ*).

The unitless estimated transmission advantage is expressed in terms of multiplicative gain in reproduction number with respect to that of the wild type, such that ℛ_variant_ = (1 + ETA) *×* ℛ_wild type_. Its calculation was made by solving the Euler-Lotka equation 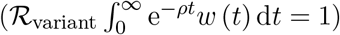 assuming a serial interval *w* following a Weibull distribution with a mean and SD of 4.8 and 2.3 days [11] and a constant ℛ_wild type_ equal to 1. The confidence interval rely on those of the estimated relative growth rate.

The estimate of the frequency of variant on Mar 12, 2021, was done by first estimating the proportion *p*_*x*_ of a given variant VX compared to the wild type (while ignoring the other variant VY) and second performing the same analysis to look at the proportion 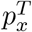 of wild type and VX compared to the whole population (VX plus VY plus wild type). The frequency of variant X was then obtained as 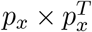.

## Supplementary figures

**Figure S1:**
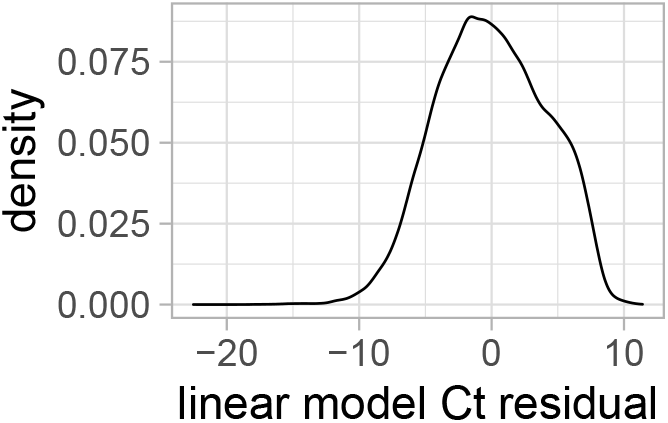
Distribution of the residual values of the multivariate linear model.

**Figure S2:**
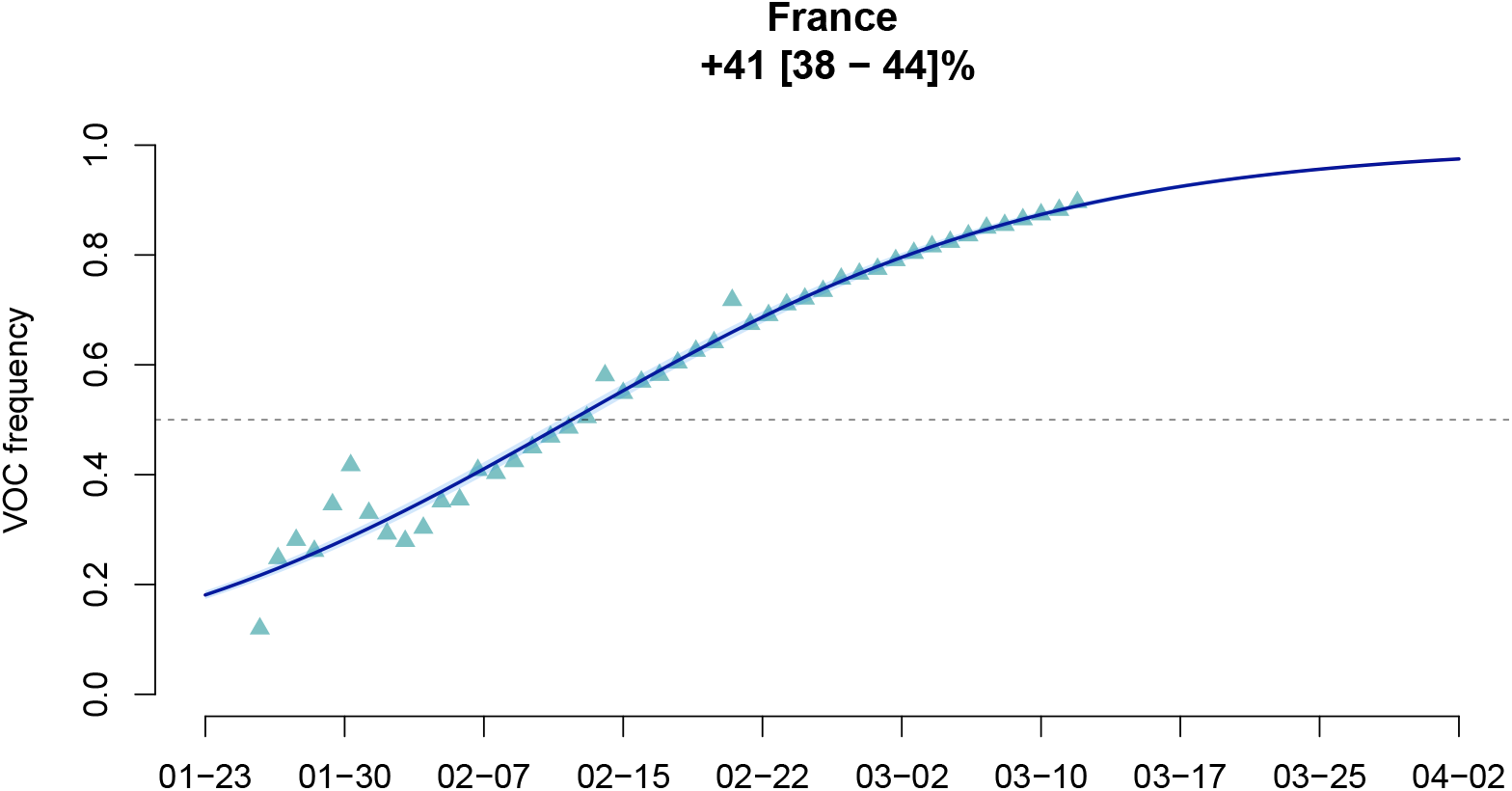
Estimating the transmission advantage of the V1 variant over the wild type strain. The dots indicate the GLM-fitted values values and the line is the output of the logisitic growth model estimation. The top figures indicate the estimated transmission advantage of the V1 variant (with respect to the wild type reproduction number) and its 95%-confidence interval. The x-axis shows the date (month-day format).

**Figure S3:**
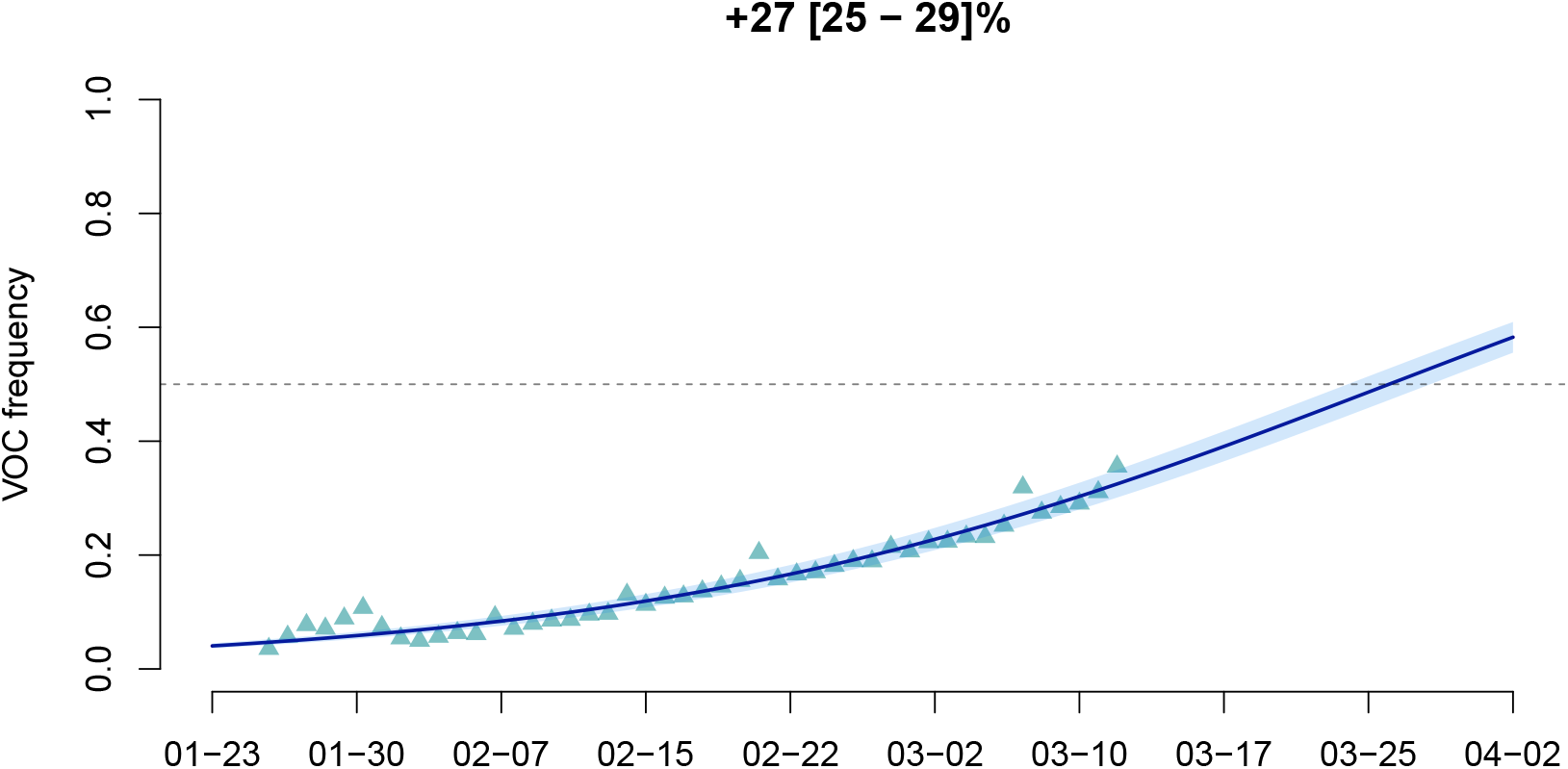
Estimating the transmission advantage of the V2 or V3 variants over the wild type strain. See Figure S2 for details.

